# A quantitative risk estimation platform for indoor aerosol transmission of COVID-19

**DOI:** 10.1101/2021.03.05.21252990

**Authors:** Hooman Parhizkar, Kevin Van Den Wymelenberg, Charles Haas, Richard Corsi

## Abstract

Aerosol transmission has played a significant role in the transmission of COVID-19 disease worldwide. We developed a COVID-19 aerosol transmission risk estimation platform to better understand how key parameters associated with indoor spaces and infector emissions affect inhaled deposited dose of aerosol particles that convey the SARS-CoV-2 virus. The model calculates the concentration of size-resolved, virus-laden aerosol particles in well-mixed indoor air challenged by emissions from an index case(s). The model uses a mechanistic approach, accounting for particle emission dynamics, particle deposition to indoor surfaces, ventilation rate, and single-zone filtration. The novelty of this model relates to the concept of “inhaled & deposited dose” in the respiratory system of receptors linked to a dose-response curve for human coronavirus HCoV-229E. We estimated the volume of inhaled & deposited dose of particles in the 0.5 to 4 μm range expressed in picoliters (pL) in a well-documented COVID-19 outbreak in restaurant X in Guangzhou China. We anchored the attack rate with the dose-response curve of HCoV-229E which provides a preliminary estimate of the average SARS-CoV-2 dose per person, expressed in plaque forming units (PFUs). For a reasonable emission scenario, we estimate approximately three PFU per pL deposited, yielding roughly 10 PFUs deposited in the respiratory system of those infected in Restaurant X. To explore the platform’s utility, we tested the model with four COVID-19 outbreaks. The risk estimates from the model fit reasonably well with the reported number of confirmed cases given available metadata from the outbreaks and uncertainties associated with model assumptions.

**Practical Implications:** The model described in this paper is more mechanistic in nature than standard probabilistic models that fail to account for particle deposition to indoor materials, filtration, and deposition of particles in the respiratory system of receptors. As such, it provides added insights into how building-related factors affect relative infection risk associated with inhaled deposited dose. An online version of this mechanistic aerosol risk estimation platform is available at Safeairspaces.com. Importantly, the modular nature of this approach allows for easy updates when new information is available regarding dose-response relationships for SARS-CoV-2 or its variants.

## 1. Introduction

Globally as of March 4 2021, more than 2.5 M deaths among 114 M confirmed cases have made COVID-19 one of the most severe diseases in history (“WHO Coronavirus Disease (COVID-19) Dashboard,” 2020). There have been many debates around the proportional routes of human-human transmission caused by large droplets, e.g., greater than 100 microns, and smaller aerosol particles that remain infectious both on surfaces and air (Liu et al., 2020). Aerosols consist of particles less than 100 *μm* in diameter that follow airflow streamlines among which smaller diameters, e.g., < 5 *μm*, can readily penetrate airways all the way down to the alveolar region where gas exchange takes place between the air and blood (Tellier, Li, Cowling, & Tang, 2019). Aerosol transmission has been implicated in several large COVID-19 outbreaks, also called “superspreading events” (Hamner et al., 2020; Hwang, Chang, Bumjo, & Heo, 2020; Khanh et al., 2020; Lu et al., 2020; Shen et al., 2020). Among the community outbreaks with well-established environmental and epidemiological analyses, there is evidence that COVID-19 may be transmitted at distances greater than two meters and may be the primary route is some COVID-19 outbreaks (Hwang et al., 2020; Kwon et al., 2020; Nissen et al., 2020). After many months of not doing so, the Centers for Disease Control (CDC) in the US indicated that aerosol transmission is believed to be a primary mode of transmission infection (CDC, 2020). The WHO still prioritizes other modes of transmission but recognizes the importance of ventilation, which primarily influences levels of aerosol particles in indoor air (WHO, 2020). Several researchers have recommended indoor air mitigation strategies for COVID-19 (Dietz et al., 2020; Morawska & Milton, 2020). Estimating infection risk by aerosol transmission and understanding the interplay of key variables that implicate the inhaled deposited dose of particles containing SARS-CoV-2 virions is important to help reduce transmission risk (Hadei, Hopke, Jonidi, & Shahsavani, 2020).

Models for airborne infectious disease transmission often rely on a quanta generation rate, which is back-calculated based on outbreaks with sufficient metadata for modeling the event (Riley, Murphy, & Riley, 1978). While this approach has been widely used, the mechanistic behavior of the environmental accumulation, fate and control of virus-laden aerosol particles is limited and largely lumped into empirically derived quanta generation rates. Such models also fail to differentiate the dynamics of different particle sizes, emission modes, and rebreathed respiratory system deposition.

In this paper we present a model based on quantitative microbial risk assessment (QMRA) for transmission of COVID-19 by aerosols in the far field. The model employs an aerosol number balance in a well-mixed indoor space with one or more infectors, mechanistic and size-resolved particle loss mechanisms, volume deposition in the respiratory systems of susceptible receptors, and infection risk based on a corona-virus dose-response relationship for humans anchored to a well-defined outbreak case in China.

## 2. Methodology

Given current research gaps, we developed the structure for an aerosol infection transmission risk estimation platform as detailed in the following sections.

### 2.1. Time-Dependent Particle Number Concentrations

The size-resolved concentration of particles emitted by an infector in a well-mixed, single-zone, indoor space is defined by the following ordinary differential equation.

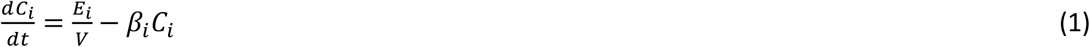

Where, *C*_*i*_ is the particle number concentration for size range i in air (particles m^−3^), E_i_ is the particle emission rate from the infector in size range i (particles h^−1^), V is the volume of the indoor space (m^3^), β_i_ is a particle removal constant (h^−1^), and t is time (h).

Particle emissions are assumed to occur from three modes as defined by Equation 2.

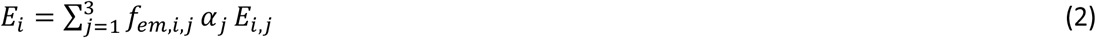

Where, counter and subscript j correspond to mode of emissions (1 = breathing, 2 = speaking, 3 = coughing), E_i,j_ is size (i) specific emission rate for breathing and speaking (particles h^−1^) and size-specific particles per cough, f_em,i,j_ is the fractional reduction in particle emissions in size range i for emission mode j as a result of the infector wearing a mask (-) (f_em,i,j_ = 0 with no mask), and α_j_ is the fraction of time exhaling (j = 1) or speaking (j = 2), or the frequency of coughing (coughs h^−1^) (j = 3). Values of f_em,i,j_ have recently been published for several different types of mask materials, assuming a good fit to face (Drewnick et al., 2021; Gandhi & Marr, 2020; Leung et al., 2020; L. Li, Niu, & Zhu, 2020; Milton, Fabian, Cowling, Grantham, & McDevitt, 2013).

We treat size-resolved particle distributions as being after the rapid evaporation process of the volatile fraction of particles. This process takes place over time scales of seconds, e.g., much shorter than typical exposure events (Chaudhuri, Basu, & Saha, 2020). Emissions are taken as time-averaged over the course of an exposure event, effectively “smoothing” intermittent emissions, e.g., coughs, and treating emissions for each particle size range as constant.

The particle removal constant, β_i_, involves four terms as described by Equation 3.

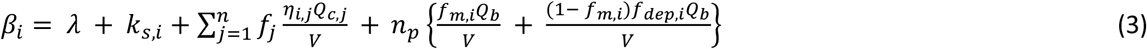

Each term in Equation 3 has units of inverse time, here taken to be h^−1^, and is assumed to be constant or time averaged (e.g., term 2) over the exposure period. The first term on the right-hand side is the air exchange rate for the indoor space (h^−1^), taken to be the volumetric flow rate of outdoor air into the indoor space (m^3^ .h^−1^) normalized by V (m^3^). Values of λ can vary in the indoor space over time and significantly between buildings. However, reasonable bounds can be placed on λ based on previous studies for residential (e.g., (Murray & Burmaster, 1995) and commercial (e.g., (Persily, Gorfain, & Brunner, 2006) buildings, standards, design, and measurements (e.g., ANSI/ASHRAE Standard 62.1-2019).

The second term, k_s,i_, is the particle decay rate due to deposition to indoor surfaces (h^−1^) for particles in size range i. The value of k_s,i_ is a function of particle size, surface-to-volume ratio in an indoor space, mixing conditions in bulk room air, air speeds over materials, and the nature of materials in the indoor space. Values of k_s,i_ for various conditions based on theory, chamber, and field experiments have been reported (Hussein & Kulmala, 2008; K. Lai & Nazaroff, 2000).

The third term corresponds to removal of particles in size range i due to engineering control devices, e.g., filtration in a mechanical system or a portable air cleaner. The parameter f_j_ is the fraction of time that air flows through device j (-), η_i,j_ is the fractional removal of particles in size range i that are removed during flow through the control device (-), and Q_c,j_ is the volumetric flow rate of air through the control device (m^3^ h^−1^). The value of each parameter in this term are system-specific. The product η_i,j_ × Q_c,j_ for a portable air cleaner is referred to as a clean air delivery rate (CADR) and is often reported in standard cubic feet per minute (scfm) based on certified testing using smoke, pollen, and dust. A range of values have been published in the literature for portable HEPA air cleaners and ion generators, the latter of which typically have much lower values of CADR (Waring, Siegel, & Corsi, 2008).

The fourth term corresponds to particle removal on the mask and in the respiratory system of the n_p_ occupants in the indoor space. The parameter f_m,i_ is the fractional removal of particles in size range I by a receptor’s mask (f_m,i_ = 0 for no mask), Q_b_ is the respiratory volume intake of occupants in the indoor space (m^3^ h^−1^), and f_dep,i_ is the fractional deposition of particles in size range i in the respiratory system of each occupant (Hinds, 2012). Values of f_m,i_ have recently been published for a wide range of mask materials, assuming a good fit to face (Drewnick et al., 2021; Gandhi & Marr, 2020; Leung et al., 2020; L. Li et al., 2020; Milton et al., 2013). Values of Q_b_ can vary by over an order of magnitude depending on body size and level of activity, e.g., rest versus heavy aerobic exercise (Epa & ORD, 2015). Values of f_dep,i_ varies by particle size, mode of breathing (mouth versus nose), and level of activity. Several different models have been developed for estimating f_dep,i_ explicitly or by computational fluid dynamics (Guha, Hariharan, & Myers, 2014; Sturm, 2016).

Separation and integration of Equation 1 yields Equation 4.

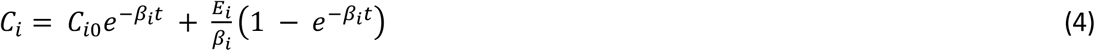

Where, C_i0_ is the initial number concentration of particles in size range i in air at the start of the exposure period (particles m^−3^), t is time (h), and all other variables are as described previously.

#### 1.1.1. Deposition of Particles in the Respiratory System

The number of particles of a specific size range deposited in three regions of a receptor’s respiratory system is estimated by Equation 5.

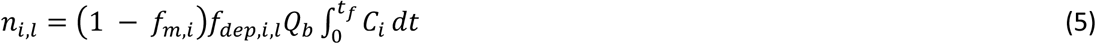

Where, n_i,l_ is the number of particles in size range i that deposit in respiratory region l (l = 1 extrathoracic region, 2 = tracheobronchial region, 3 = alveolar region), f_dep,i,l_ is the fractional deposition of particles in size range i in region l of the respiratory system (Hinds, 2012), and all other variables are as defined previously. The integration of C_i_ is taken from initial exposure time (time = 0 h) to final exposure time t_f_ (h).

The volume of particles of a specific size range deposited in the aforementioned regions of a receptor’s respiratory system is estimated by Equation 6.

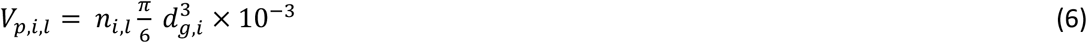

Where, V_p,i,l_ is the volume of particles in size range i that deposit in respiratory region l (pL), and d_g,i_ is the geometric mean diameter based on end points of size bin i (μm). The total volume of particles across all particle sizes and regions of the respiratory system is determined by Equation 7.

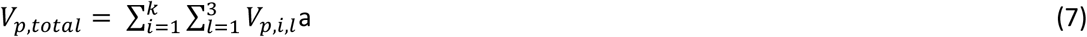

Where, V_p,total_ is total volume of all particles deposited in the receptor’s respiratory system summed over all k particle size ranges and three respiratory regions (pL).

Many of the parameters described above are building or indoor space specific and can be easily varied in the model, e.g., to model known outbreaks or to draw comparisons between different hypothetical scenarios. The model can incorporate emissions and particle size distributions reported in the published literature or as new data become available. For the purpose of examples presented in this paper, we use data from the study of size-resolved particle emissions associated with speaking (Asadi et al., 2019), and another study of size-resolved particle emissions associated coughing (Lindsley et al., 2012), and an approximation that emissions from breathing are 10% of those from speaking with median amplitude. We further assume, for this analysis, the possibility of a “high emitter” and “low emitter” of aerosol particles. Profiles for each described below:

High emitter:

- Coughs eight times per hour
- Each cough emits 54,000 particles
- Cough particle sizes are defined by case #8 in Lindsley et al. (2012)
- Emitter spends 20% of the event time speaking at an elevated amplitude with size-specific emissions as per Asadi et al. (2019).

Low emitter:

- Same as high emitter but without cough and median amplitude.

### 2.2. Model Application to Guangzhou Restaurant Outbreak

Ten persons from three families (families A–C) who had dined at the same restaurant (Restaurant X) in Guangzhou, China, became infected with COVID-19 (Lu et al., 2020). A layout of the restaurant and infection zone is provided in Figure 1. Family A traveled from Wuhan and arrived in Guangzhou. The next day the index case (Case A1) ate lunch with three other family members (A2–A4) at Restaurant X. Two other families, B and C, sat at neighboring tables at the same restaurant. Later that day, Case A1 experienced the onset of fever and cough and went to the hospital. Several days later a total of nine others (4 members of family A, 3 members of family B, and 2 members of family C) had become ill with COVID-19. The only known source of exposure for the affected persons in families B and C was Case A1 at the restaurant.

**Figure 1.**
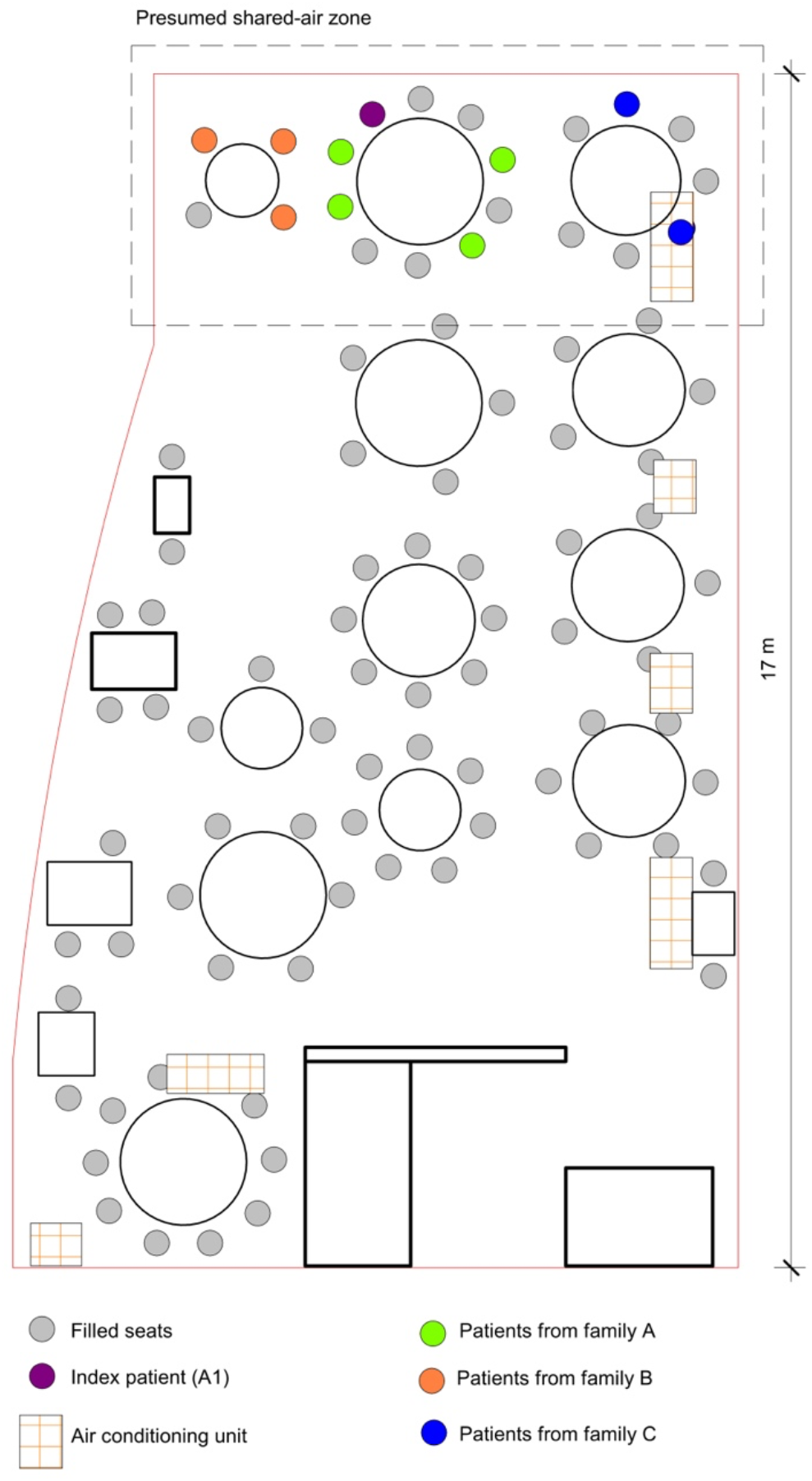
*arrangement of restaurant tables and air conditioning airflow at site of outbreak of 2019 novel coronavirus disease, Guangzhou, China, 2020, following* (Y. Li et al., 2020)

The model was applied to this outbreak in Restaurant X in Guangzhou, China with significant metadata available (Y. Li et al., 2020; Lu et al., 2020; Moses, Gonzalez-Rothi, & Schmidt, 2020). A closed-circuit video was used to determine the position of a single infector (index case) as well as receptors who became infected following the event and exposure times. An inert tracer gas was used to study air flow patterns and to quantify air exchange rates.

For the analysis described herein only respirable particles between 0.5 and 4 μm with size ranges (bins) of 0.5 μm were used to estimate total volume of particles deposited in the respiratory systems of those infected (Equation 8). The upper bound of this range can be easily extended in the model. The infector was assumed to speak with high amplitude 20% of the time and cough with a frequency of 8 coughs/hr.

Receptors were assumed to have a respiratory minute volume of 0.6 m^3^/hr and were nose breathers. The exposure period was taken to be approximately 1.25 hours. Particle deposition in the respiratory system for nose breathers was based on the ICRP 66 model (“Human Respiratory Tract Model for Radiological Protection. A Report of a Task Group of the International Commission on Radiological Protection,” 1994).

Virus-laden aerosol particles were assumed to be constrained to an infection zone with recirculated air over three tables, including that where the infector was seated. This is a conservative assumption, overestimating the inhaled volume by neglecting dispersion out of the recirculated infection zone. Particle deposition to surfaces was based on values of k_s,i_ as reported by Hussein and Kulmala (2008), but was generally small across particle sizes considered.

Parameters used to simulate the outbreak in Restaurant X are shown in Table 1. We acknowledge that the aerosol particle emission profile for the infector is hypothetical and based solely on reasonable values in the literature. Other parameters in Table 1 are those reported for the infection zone in Restaurant X.

**Table 1.** Guangzhou Restaurant X risk estimation input assumptions

| Guangzhou restaurant physical parameters | | Emission parameters (0.5-4 $\mu\text{m}$) | |
| --- | --- | --- | --- |
| Occupants (#) | 21 | Cough (particles/cough) | 54,000 |
| **Time of event (hr)** | 1.25 | **# coughs/hour** | 8 |
| **Floor area (m<sup>2</sup>)** | 35 | **Speak (total particles/hr)** | 360,000 |
| **Ceiling height (m)** | 3.14 | **Fractional time speaking** | 0.2 |
| **Outdoor Air Supply (m<sup>3</sup>/hr)** | 61.5 | **Breathing (particles/hr)** | 36,000 |
| **CADR - filtration (m<sup>3</sup>/hr)** | 0 | **Fractional time not speaking** | 0.8 |
| **Speak multiplier** | 1.5 |  |  |

An additional speak multiplier function was enabled to count for conditions in which index and susceptible patients’ activities involve higher metabolic activities such as speaking loudly, which multiplies the initial 360,000 particles/h^−1^ to corresponding values. A model simulation for total particle concentration in the zone of infection in Restaurant X is shown in Figure 2. The peak concentration for particles in the size range of 0.5 to 4 μm is 3,800 m^−3^ at the time when the infector left the restaurant. steady-state condition was not achieved during the infection event, and the average concentration of particles is estimated to be 2236 m^−3^.

**Figure 2.**
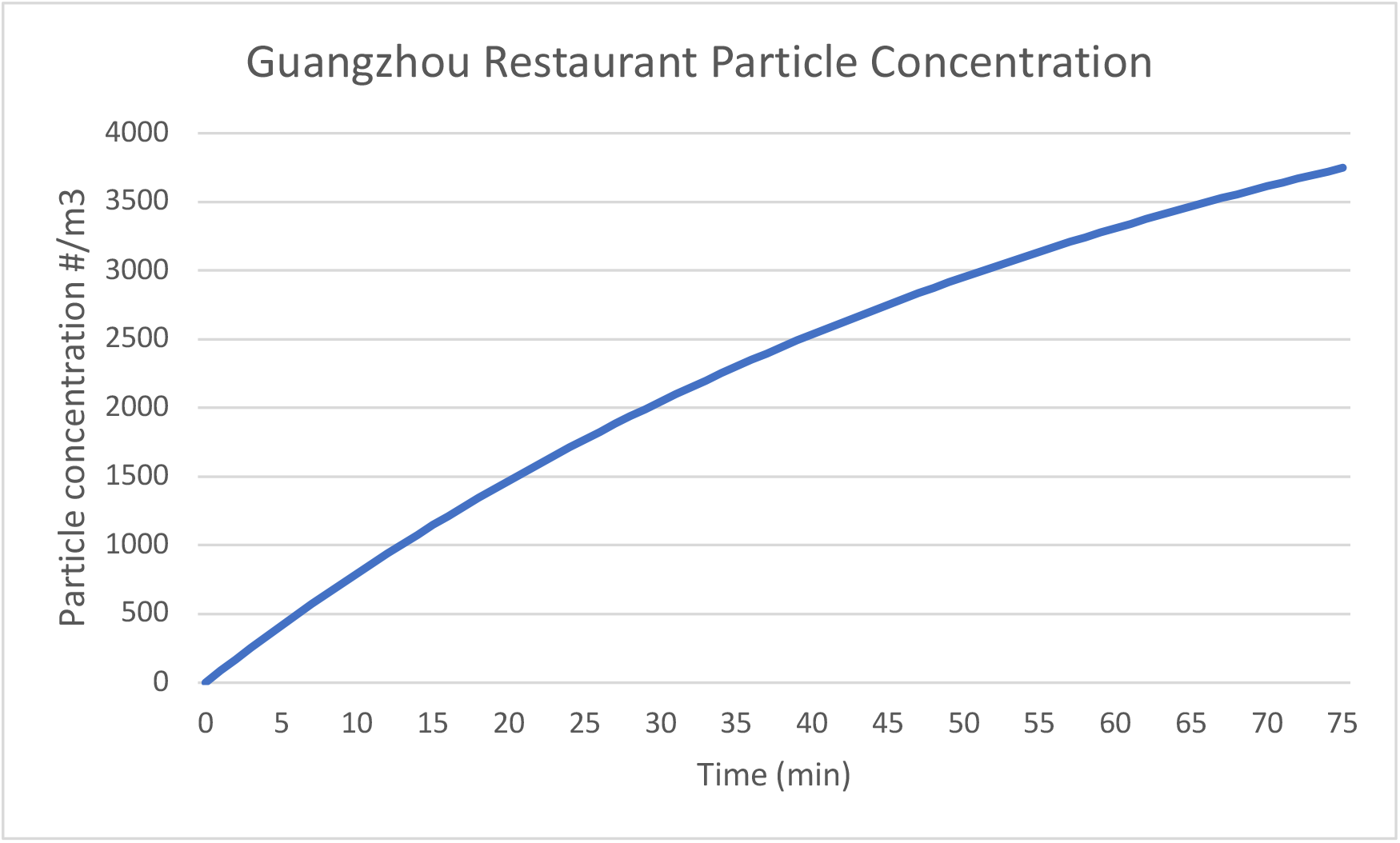
Total particle concentration in restaurant zone during time infector is in the space.

**Figure 3.**
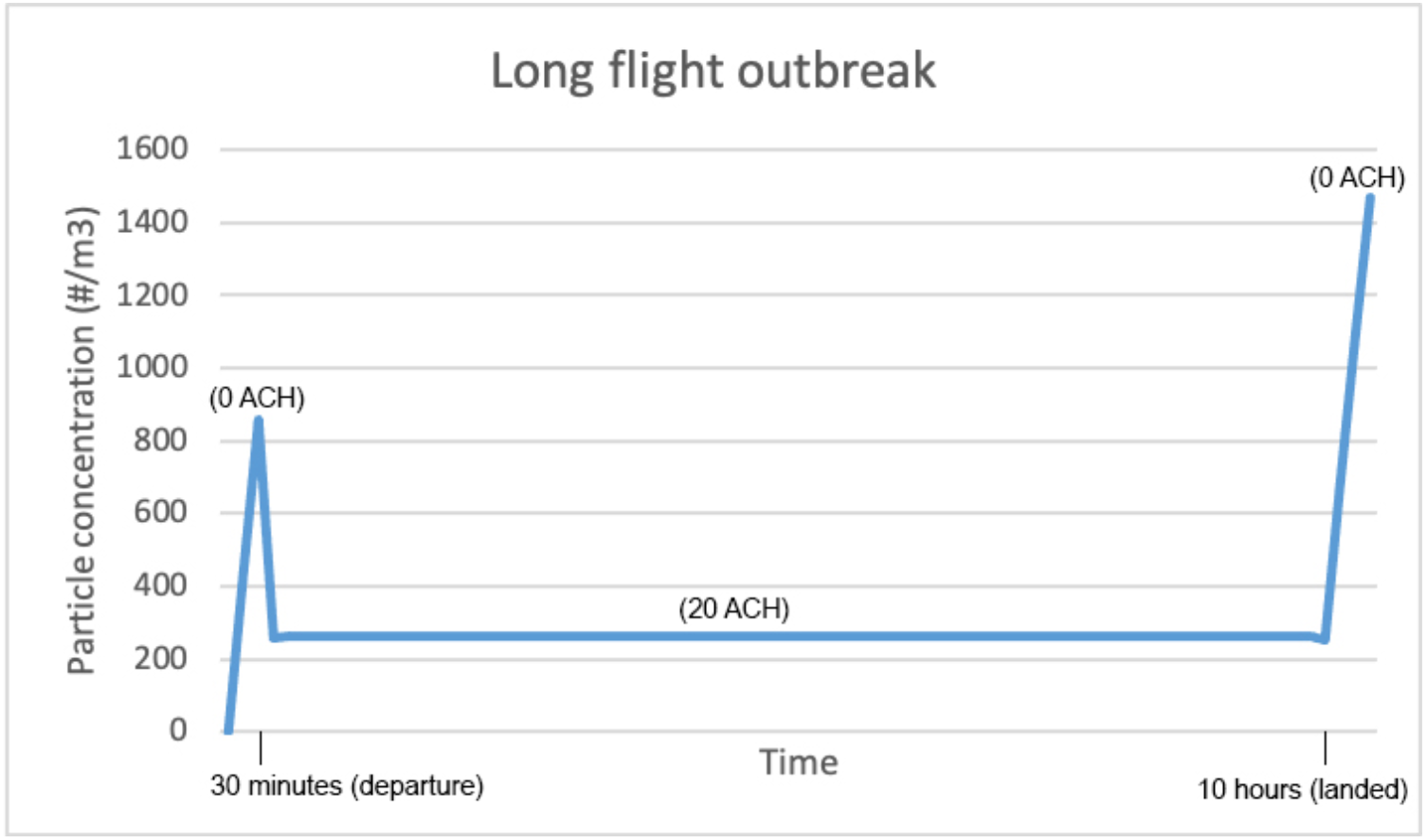
Long flight outbreak emission over time

For the parameters used in this simulation we estimated that, on average, each individual in the space had 3.6 picoliters (pL) of aerosol particles in the 0.5 to 4 μm range deposited in their respiratory system from a high emitter during the infection event. The particle volume was 0.58 pL, 0.25 pL, and 2.78 pL deposited in alveolar, trachea bronchia, and head airways, respectively. Based on a number of simulations with a range of reported particle emission rates for coughing, speaking and breathing, it seems reasonable that actual volume deposited was in the range of 0.5 to 10 pL. For the remainder of this analysis, we use 3.6 pL for purposes of illustration and comparison.

Once the (average) dose to a receptor is estimated, a dose response curve can be used to assess the risk. At present, no data are available to construct a dose-response curve for SARS-CoV-2. However prior work has shown that dose-responses for other coronaviruses obey an exponential relationship as described by Equation 8 (Watanabe, Bartrand, Weir, Omura, & Haas, 2010).

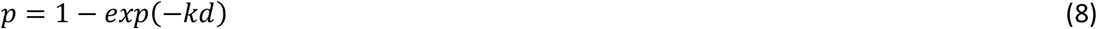

Where p is the proportion of exposed individuals adversely affected (probability of infection), and d is the average viral dose that those individuals were exposed to in plaque forming units (PFU). In the absence of data for SARS-CoV-2, the epidemiologic data from the Guangzhou outbreak is used to anchor the dose response relationship in the same sense that has been used in food microbial risk assessment (Miliotis, Dennis, Buchanan, & Potter, 2008). In this approach, the parameter “k” is calibrated to match the attack rate from the exposure observed in the outbreak.

For the outbreak in Restaurant X, there are at least two ways to consider the infection rate, and therefore the response in the dose-response model. One is that the index emitter’s particles resulted in 9 people out of 20 becoming infected with COVID-19, yielding an attack rate of 45%. Another is that 5 out of 11 people that were not at the same table as the index individual were infected via aerosols in the space during the event. There are two justifications for the latter, one being that the infected individual was travelling with two family members and may have spread the virus to them shortly before or after the event at the restaurant, and the other is that the other non-family member at that same table that became infected may have been infected via large droplets or direct contact with the index individual rather than via aerosols. Nonetheless, 5 infected of 11 susceptible individuals results in a similar attack rate of 45%. Therefore, we calculated the probability risk of 45% for this case study.

Using a dose-response model developed by Haas (2020) we calculated a dose of 11 PFUs to yield the observed 45% attack rate in Restaurant X. There is some evidence that this magnitude of viral dose may yield infection based upon previous influenza research (Alford, Kasel, Gerone, & Knight, 1966). Assuming a volume deposition of 3.6 pL per person within the zone of interest yields an average of ∼3 PFUs/pL deposited. We anchored our calculations to the Haas dose-response curve using 3 PFUs/pL to extrapolate the risk probability of our SARS-CoV-2 platform with a dose in pL. While a broader range of particle sizes can be employed in the model, for this illustrative analysis we assume that the model scales linearly with similar infectious viral load per pL for all particle sizes.

## 3. Results and Discussion

Rather than presenting results per se, we have organized a series of case studies into a discussion with two sections. This is meant to help potential users of the screening model platform to develop a practical understanding of its capabilities and limitations. The section uses the platform to simulate four well-known COVID-19 outbreaks as a means to explore its utility and generalizability.

### 3.1. Case 1. Bus ride in Eastern China

People who rode one bus to a worship event and back, in which there was at least one confirmed COVID-19 case, had a statistically significant higher risk of SARS-CoV-2 infection than individuals who rode a different bus to the same event. In the first bus, 23 out of 68 passengers tested positive for COVID-19 while none of the passengers in the second bus were diagnosed with COVID-19 (Shen et al., 2020). Table 2 shows summarizes the inputs used in the risk estimation platform.

**Table 2.** Bus riders in eastern China assumption

| Bus riders in Eastern China |  |  |  |
| --- | --- | --- | --- |
| Occupants (#) | 68 | Outdoor Air Supply (m<sup>3</sup>/hr) | 3.3 -9.19 |
| Time of event (hr) | 1.66 | Filtration CADR (m<sup>3</sup>/h) | 0 |
| Volume (m<sup>3</sup>) | 80 | Fractional time speak | 0.2 |
| Speak multiplier | 1 | Coughs h<sup>-1</sup> | 8 |
| High emitter | 1 | Low emitter | 0 |

The air exchange rate for the bus was not published. Using 4 air changes h^−1^ in the screening model described herein yields the observed 23 infections and 34% infection probability. We were only able to find one published study for which the air exchange rate for a bus was reported. Previously, researchers used sulfur hexafluoride release and decay and reported air exchange rates of 2.6 to 4.6 h^−1^ for a traveling school bus on its normal route (Rim, Siegel, Spinhirne, Webb, & McDonald-Buller, 2008). This range bounds the air exchange rate of 4 h-1 that yields a model result consistent with disease cases in the outbreak.

### 3.2. Case 2. Two choir rehearsals in Skagit Valley

Another outbreak event occurred on the evening of March 10, 2020, in which 32 out of 61 members that attended a weekly rehearsal were confirmed positive for COVID-19 and another 20 were symptomatic but not tested or confirmed positive (Hamner et al., 2020; Miller et al., 2020). It is not clear whether all 32 cases (or 52 including unconfirmed and symptomatic) acquired the infection during the event on March 10^th^ or whether some may have acquired the infection during the previous weekly practice on March 3^rd^. Importantly, three cases were confirmed in less than 24 hours and five more in less than 48 hours after the March 10^th^ event, fairly short periods for symptom onset or positive test, and suggests that some may have been infected during the March 3^rd^ practice or elsewhere. Therefore, it is conceivable, but not shown, that some or all of these eight individuals could have also been contributing infectors during the March 10^th^ event.

In this outbreak, the major unknown variables are the air exchange rate, how many choir members were emitting viral particles during the infection event, and which susceptible individuals were in contact with potential emitters for what durations during three sub-events on March 10^th^. The observed attack rate of the event, ranging from 53.3% to 86.7% (32-52 infected) is also in question given the potential number of additionally infected but pre- or asymptomatic choir members. Ventilation rates were estimated to be 0.3 - 1h^−1^ based on environmental heat balance estimates (Miller et al., 2020). Therefore, we simulated this case in using the risk estimation platform with a range of high and low emitters, as well as three air exchange rates. Our assumptions for evaluating this case are outlined in Table 3.

**Table 3.**
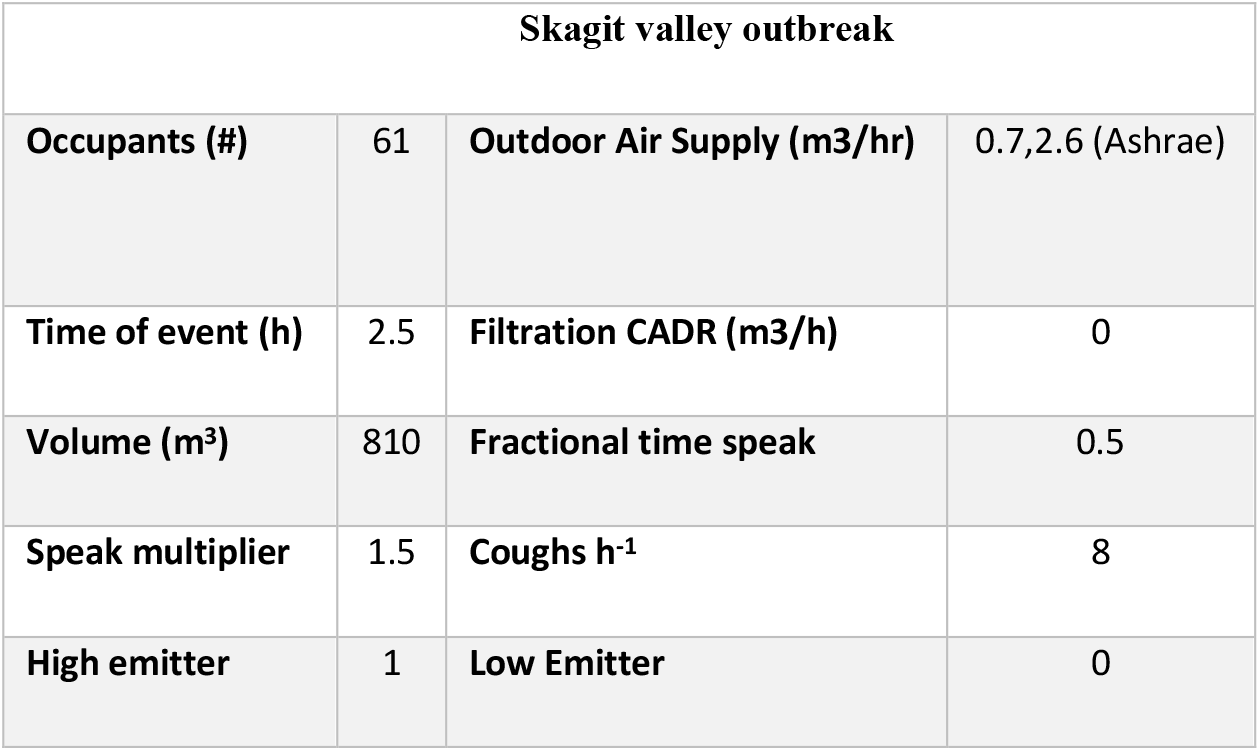
Skagit Valley choir assumptions

| Skagit valley outbreak |  |  |  |
| --- | --- | --- | --- |
| **Occupants (#)** | 61 | **Outdoor Air Supply (m3/hr)** | 0.7,2.6 (Ashrae) |
| **Time of event (h)** | 2.5 | **Filtration CADR (m3/h)** | 0 |
| **Volume (m<sup>3</sup>)** | 810 | **Fractional time speak** | 0.5 |
| **Speak multiplier** | 1.5 | **Coughs h<sup>-1</sup>** | 8 |
| **High emitter** | 1 | **Low Emitter** | 0 |

According to the number of potential infectors in the space, we simulated this outbreak as the following three scenarios:

#### (a) 1 high emitter (the index case)

According to this scenario for a ventilation rate of 0.7 h^−1^, each individual would have received a deposited respiratory dose of 2 pL. Our estimated simulation and the presence of one super spreader (the index case) would result in an estimated risk of 29%, yielding 18 people being infected during the event. It is possible that emissions from the single infector were higher than our assumed index case, the air exchange rate much lower, or that additional infectors were preset during the infection event.

#### (b) 1 high emitter and 7 low emitters in the space (including those who tested positive on March 11 & 12)

Assuming that COVID-19 qRT-PCR tests after 1-2 days of exposure are not likely to produce positive results (due to virus incubation period), there could have been more than one index emitter in the space, with up to seven additional pre-symptomatic emitters in the rehearsal (Cevik et al., 2021). Therefore, we also simulated the outbreak with 1 high emitter and 7 low emitters (individuals who were confirmed positive on March 11 and March 12). With these inputs, the model estimates an infection probability of 87%, yielding 53 additional infections (for a total of 61), which is more than the number of reported (confirmed and suspected) cases.

#### c) 4 high emitters in the space (including those who tested positive on March 11)

In this case, the model estimates 46 additional infected individuals, for a total of 50, representing an infection probability of 75%.

There are a lot of unknowns regarding this Skagit Valley Choir outbreak and the primary intent of this analysis is to show how the model can be used to rapidly assess a range of scenarios or to potentially calibrate one scenario.

### 3.3. Case 3. A long flight from London to Hanoi

Air travel is commonly judged to have low risk of transmitting SARS-CoV-2 through cabin air due to high ventilation rates and recirculation through HEPA filtration (Richard, 2020). However, it is conceivable that even relatively low particle concentrations associated with an infector can become an infection transmission concern on long flights (Gendreau, 2010).

A COVID-19 outbreak was reported to have been associated with a 10-hour flight from London, UK, to Hanoi, Vietnam, in early March 2020 (Khanh et al., 2020). There were 16 crew members and 201 passengers on board. The index case was identified as having symptoms including sore throat and cough that began 3 days prior to the flight. She was confirmed positive after the flight through qRT-PCR test (Khanh et al., 2020). Among passengers who remained in Vietnam and were traced (all except 30 passengers), 16 positive cases were detected, including 12 (80%) which had travelled in business class with the index case. Additionally, one case was in economy class, and one case was among flight attendants that likely traversed between cabins. The business class section is spatially divided from the economy class with kitchens and bathrooms, and therefore we focused analyses on this discreet zone. We considered the impact of lower ventilation rates (approximately 0 h^−1^) in the airplane cabin while the plane was on the ground, taxiing on the runway, and at the gate for boarding and deplaning. Then, we estimated a higher air exchange rate of 20 h^−1^ supplied in the cabin during the flight, whereby 10 ACH of outside air is provided (representing 50% outside air fraction), and the equivalent of 10 ACH of additional clean air delivery (1650 m^3^/h) is provided via central HEPA filtration. Business class passengers typically board before the main cabin and therefore experience lower airflow rates for longer periods of time preflight, thus increasing the initial aerosol concentration at the beginning of the event. Figure 4 and Table 4 outline the parameters used in the simulation.

**Figure 4.**
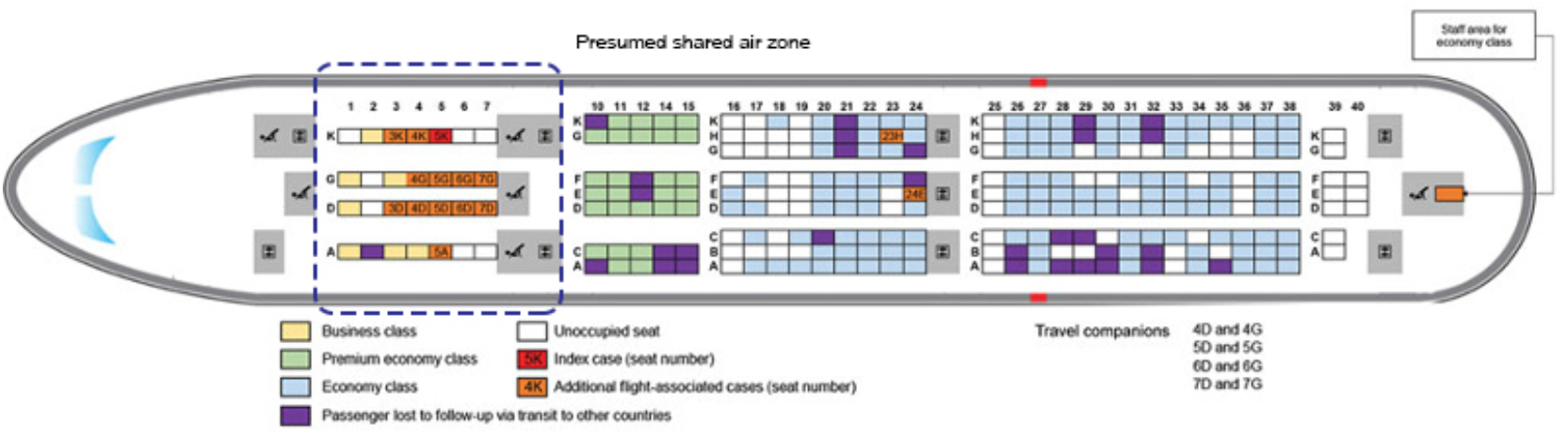
Seating location of passengers on Vietnam Airlines flight 54 from London, UK, to Hanoi, Vietnam, on March 2, 2020 following (Khanh et al., 2020)

**Table 4.** Long flight outbreak assumption

| Long flight outbreak |  |  |  |
| --- | --- | --- | --- |
| Occupants (#) | 21 | Outdoor Air Supply during flight (m<sup>3</sup>/hr) | 10 ACH |
| Time of event (hr) | 0.5 (boarding)<br>9.5 (flight)<br>0.5 (departure) | Outdoor Air Supply during boarding & departure (m<sup>3</sup>/h) | 0 |
| Volume (m<sup>3</sup>) | 165 m<sup>3</sup> | Filtration CADR (m<sup>3</sup>/h) | 1650 |
| Speak multiplier | 1 | Coughs h<sup>-1</sup> | 8 |
| High emitter | 1 | Low emitter | 0 |

We created a digital model of the interior business class section of the AIRBUS-A350-900 aircraft and estimated 165 m^3^ as the volume of the zone. Since the index case had symptoms during the flight, we simulated this event with a high emitter (as explained in methodology section). According to the simulation with these parameters, each passenger was likely to have a respiratory deposited dose of 2.8 pL, which yields an estimated 37% infection probability and 8 additional passengers infected, close to the 10 infections reported. Ten infections (49% attack rate) would require 4 pL of deposited dose from either a higher emitter (15 more coughs h^−1^) or an air exchange rate of 12 h^−1^ during the in-flight period, both of which are reasonable values.

It is possible that large droplet, contact transmission, or concentrated aerosol plume transmission occurred between the index case and nearby passengers. According to the seating positions of the index case and the additional confirmed cases during the flight (Khanh et al., 2020), some passengers (especially those sitting in 3K and 4K) may have been infected through large droplets or concentrated aerosol plume emitted from the index case coughing. However, infected passengers who were positioned in rows G, D, and A are more likely to have been exposed to the virus through only aerosol transmission. It is also possible that some transmission may have occurred prior to the flight, in the airport, boarding areas, or while boarding or deplaning. Still, given these dynamics and possible alternate scenarios, the model reasonably estimates the number of reported cases likely to have occurred via aerosol transmission.

### 3.4. Case 4. Transmission of SARS-CoV-2 by direct airflow in restaurant X in South Korea

According to a well-characterized epidemiological study for an event on June 17, 2020, a confirmed COVID-19 case was identified to have been infected in a restaurant in Jeonju, South Korea where indoor air circulation may have transmitted SARS-CoV-2 virus 6.5 m away from the infector (Kwon et al., 2020). Case A and C were reported to be infected after 5 minutes and 20 minutes, respectively. According to floor plans (Figure 5A), these cases were downstream of aerosols likely to contain SARS-CoV-2 virions expired by Case B and potentially Case D. Case D was travelling with index Case B but didn’t have symptoms during the event. The restaurant had a total floor area of 97 m^2^, and reportedly had no windows or ventilation systems (Kwon et al. 2020). For event evaluation purposes, we assumed 77 m^2^ of this space (excluding the kitchen area) is a shared-air zone and was used as an input to the risk estimation platform. Note that the available evidence suggests that this space may not have represented a will-mixed air volume, thus presents a challenging case to the risk model. We simulated this event with one high emitter in one scenario and one high emitter and one low emitter (including case D) in an alternate scenario. The event duration is also unique in that there are important time overlaps between cases as outlined in Figure 5B, presenting another challenge to the risk model. Case A only overlapped with Cases B & D for 5 minutes. Case C overlapped for 20 minutes, but also remained in the space for another 27 minutes after Cases B & D departed and would still be breathing particles emitted by B & D for that duration. Thus, the particle concentration over time (#/m^3^) was expanded to account for the decay of expired particles for a period of 28 minutes after the emitter(s) had departed (Figure 5B). Therefore, the event duration was estimated to be 55 minutes (0.92 h) for the primary scenario, while other specific event duration scenarios are also described. Simulation inputs are summarized in Table 5.

**Figure 5.**
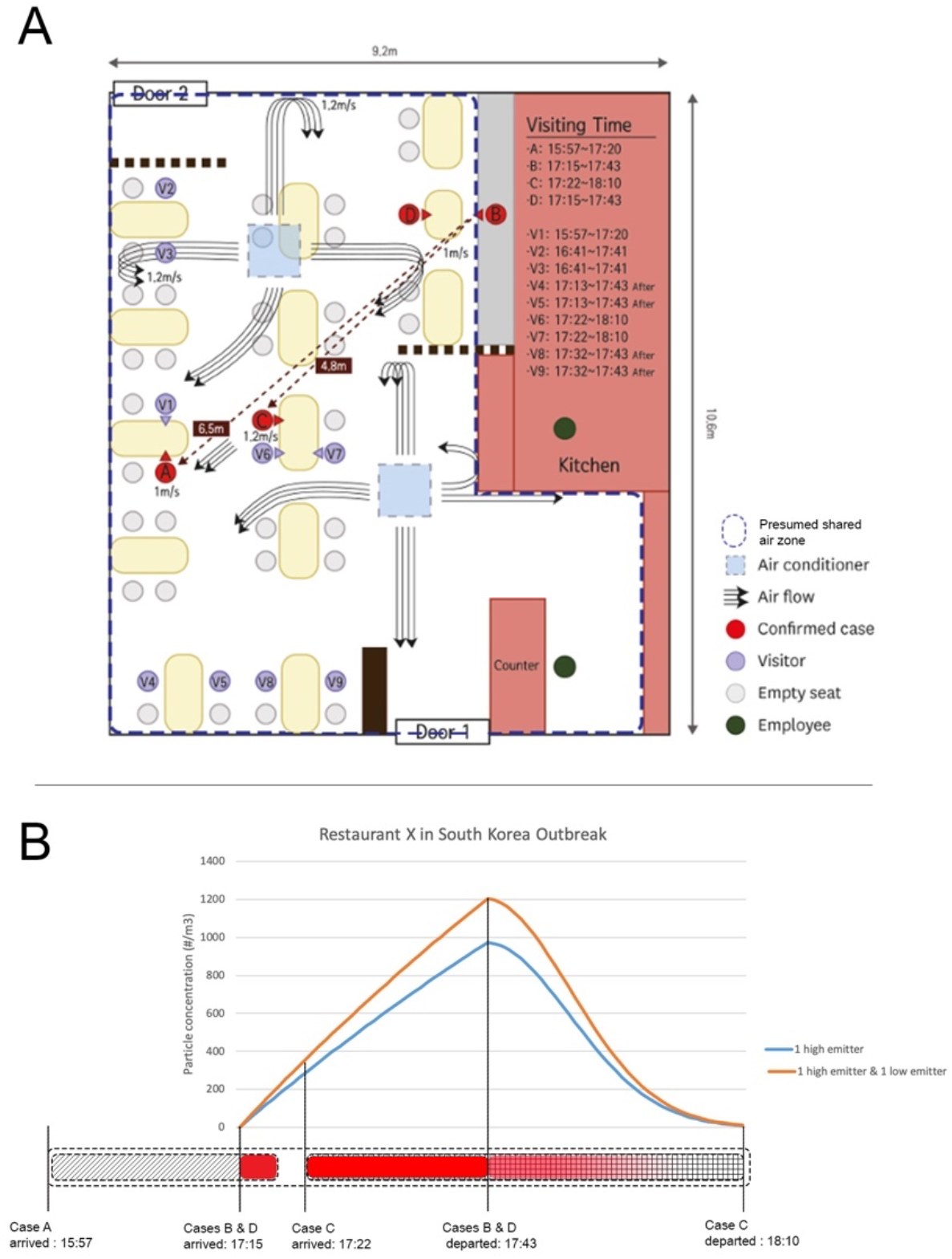
Schematic floor plan of the South Korea outbreak restaurant, following (Kwon et al., 2020)

**Table 5.** South Korea restaurant outbreak assumptions

| South Korea restaurant outbreak |  |  |  |
| --- | --- | --- | --- |
| **Occupants (#)** | 13 | **Outdoor Air Supply**<br>**(m<sup>3</sup>/hr)** | 0.2 |
| **Time of event (hr)** | 0.92 | **Filtration CADR (m<sup>3</sup>/h)** | 0 |
| **Volume (m<sup>3</sup>)** | ~185 | **Fractional time speak** | 0.2 |
| **Speak multiplier** | 1.5 | **Coughs h<sup>-1</sup>** | 8 |
| **High emitter** | 1 | **Low emitter** | 0 and 1 |

The model estimated an infection probability of 9% for the entire (55-minute) event assuming Case B was emitting aerosols into the shared-air zone for a total duration of 27 minutes, and these aerosols containing SARS-CoV-2 virions remained in the air for additional 28 minutes following a decay curve presented in Figure 5B. This 9% attack rate estimates that one person would acquire infection. Assuming that Case D contributed particles containing virions as a low (asymptomatic) emitter, the probability of infection increases to 13.5%, estimating ∼2 persons would acquire infection, similar to the 15.4% (2 infections) observed. Given what is reported in this case, it is probable that the directional airflow patterns contributed to a higher concentration of particles accumulating in the specific region within the restaurant where the rapid 5-minute exposure resulted in infection. However, despite this challenging event profile, the model reasonably estimates the event outcome.

## 4. Conclusion

There is compelling evidence that shows aerosol transmission has played an important role in the spread of COVID-19 globally. We have developed a mechanistic aerosol transmission risk estimation platform that incorporates the best available information regarding respiratory particle dynamics, viral viability, human respiratory physiology, and viral dose-response proxies, to estimate infection probability based on a list of critical inputs. A lite version of the model is available at Safeairspaces.com.

In addition to the Restaurant X outbreak in Guangzhou, China, which provided a basis for anchoring the volume of inhaled and deposited dose into probability of infection, four other well characterized outbreaks were simulated using the platform in conjunction with the best available data from epidemiological investigations. The simulations demonstrate that the risk estimation platform yields results that reasonably predict outbreak attack rates using the available information about the case and reasonable assumptions for missing information. Although it is based on several assumptions, including a dose response curve from a different Coronavirus (HCoV-229E), we believe the platform is useful now, and the mechanistic approach will rapidly accommodate updates as soon as new information becomes available, especially with regard to SARS-CoV-2 human dose-response data.

## Data Availability

This is a model for infectious disease transmission risk in indoor environments. All data used in this study were publicly available.

https://safeairspaces.com/

## Author Contributions

RC and KVDW conceived of the project scope and oversaw project and manuscript development. RC developed the initial mechanistic model. All authors collaborated to anchor the model with meta data from Restaurant X in Guangzhou China and dose-response data from HC229-E. HP and RC wrote the initial manuscript and KVDW, CH provided significant edits to the manuscript. HP and KVDW developed and RC reviewed the online user interface.

